# Characterizing Autonomic Dysfunction during Resuscitation in Sepsis using Multiscale Entropy

**DOI:** 10.64898/2026.03.04.26347662

**Authors:** Preethi Krishnan, Andrea Sikora, Brian Murray, Ayman Ali, Mihai Podgoreanu, Pulakesh Upadhyaya, Alasdair Gent, Tilendra Choudhary, Andre L. Holder, Annette Esper, Rishikesan Kamaleswaran

## Abstract

**Rationale:** Autonomic dysfunction is a hallmark of sepsis pathophysiology, yet its quantification remains challenging. Multiscale entropy (MSE) derived from heart rate variability (HRV) offers a dynamic measure of physiological complexity and may serve as a biomarker of early deterioration associated with subsequent organ failure, vasopressor escalation, or mortality.

**Objective:** To determine whether MSE computed across multiple temporal scales during the first 24 hours of Intensive Care Unit (ICU) admission is associated with short-term mortality and longer-term organ dysfunction in patients with sepsis, and whether these relationships vary across vasopressor exposure. Unlike prior studies that focused on short-term HRV metrics, we applied MSE across multiple temporal scales and incorporated these features into machine learning models to evaluate their prognostic utility in septic shock.

**Methods:** This retrospective cohort study included adult ICU sepsis patients at Emory University Hospital from January 2016 to December 2019. Of 2,076 eligible patients, 958 were propensity matched into two cohorts: fluids-only and fluids-plus-vasopressor, with norepinephrine as the primary vasopressor. High-resolution electrocardiogram (ECG) waveforms were analyzed to compute MSE across 20 temporal scales. Machine learning models using (1) MSE features alone and (2) MSE combined with demographic and vital sign data (MSE-DV) were compared against traditional HRV measures based model and severity of illness scores for predicting outcomes. Model performance was assessed using the area under the receiver operating characteristic curve (AUROC), with a primary outcome of mortality at day 7 and secondary outcome of persistent organ dysfunction at day 28.

**Results:** In the fluids-plus-vasopressor cohort, MSE-based models demonstrated superior predictive performance for 7-day mortality (AUROC 0.84) compared to severity of illness scores (AUROC 0.64). MSE-DV models also predicted organ dysfunction including 28-day renal (AUROC 0.75), neurological (AUROC 0.79), and respiratory (AUROC 0.71) dysfunction. Patients receiving second-line and third-line vasopressors and corticosteroids exhibited progressively lower MSE values, particularly at mid-range and long-range scales.

**Conclusion:** MSE features in the first 24 hours of ICU stay predict mortality and organ dysfunction with higher discrimination than traditional severity of illness scores. Future work should validate these findings, assess longitudinal MSE trends, and race-specific autonomic patterns to refine predictive models.

## Introduction

Sepsis outcomes are significantly influenced by a dysregulated host response to infection, driven by complex neuroendocrine-immune interactions.^1^ Evaluating the severity of this dysregulation is vital for prognostication. Heart rate variability (HRV) and multiscale entropy (MSE) offer quantitative measures of autonomic dysfunction, a key aspect of sepsis pathophysiology.^2–4^ Despite the potential of entropy-based approaches, prior work has often been limited by a focus on lower time scales,^4,5,6^ which may not fully capture the dynamic evolution of neuroendocrine-immune responses over time. This raises the question of whether entropy measures calculated at longer time scales can provide a more robust and relevant assessment of dysregulation and prognostication, particularly during fluid resuscitation, septic shock, and guideline-based therapy.

A significant challenge in using HRV as a reliable biomarker for predicting outcomes in septic shock is the profound influence of concurrent treatments.^7^ Vasopressors like norepinephrine directly stimulate adrenergic receptors, heavily influencing sympathetic outflow, while corticosteroids can modulate the inflammatory response which in turn affects autonomic balance.^8^ Improvements in perfusion and attenuation of the underlying sepsis process as a result of successful treatment may themselves indirectly impact HRV. These interdependent physiologic and therapeutic effects make it difficult to discern whether observed changes in HRV reflect the severity of the underlying sepsis or the influence of treatment.^9,10^ Consequently, rigorous evaluation of entropy-based metrics in sepsis has historically been limited, particularly in the context of multi-level, time-series treatment regimens and heterogenous pattern trajectories.^11^

To address these gaps, we evaluated MSE derived from high-resolution electrocardiogram (ECG) as a prognostic marker of short-term mortality and longer-term multi-organ dysfunction in patients with sepsis receiving guideline-based treatment. Building on the known sensitivity of MSE to inflammatory states and its potential to capture complex physiological dynamics, we hypothesized that MSE in the longer-range scales would serve as a robust predictor of outcomes across different treatment profiles.^12^ Because vasopressors exert substantial autonomic effects, we examined whether the relationship between MSE and outcomes varied according to vasopressor exposure. Our overarching objective was to investigate whether MSE has independent predictive power for outcomes in patients with sepsis despite the potentially confounding effects of concurrent therapies.

## Methods

This study was approved by the Institutional Review Boards (IRB) at Duke University (IRB #Pro00115678) and Emory University (CR002-STUDY00004827). All methods were performed in accordance with the ethical standards of the Helsinki Declaration of 1964. This evaluation followed Transparent Reporting of a multivariable prediction model for Individual Prognosis Or Diagnosis – Artificial Intelligence (TRIPOD-AI) reporting standards as applicable to machine learning-based prognostic modeling.^13^

### Study design

This retrospective cohort study included patients diagnosed with sepsis or septic shock. Patients admitted between January 2016 and December 2019 to an intensive care unit (ICU) at Emory Healthcare, including medical, surgical, neurocritical, transplant, and cardiac ICUs, were screened for inclusion. Eligible patients were adults aged 18 years or older who met the Sepsis-3 criteria as defined by the Third International Consensus Definitions.^14^ Patients were classified into two cohorts: a fluids-only cohort, defined by receipt of ≥30 mL/kg fluid resuscitation, and fluids-plus-vasopressor cohort, defined by receipt of ≥30 mL/kg fluid resuscitation plus first line vasopressor therapy with norepinephrine. Fluid volume for the fluids-only cohort was calculated over the 12 hours before and after meeting Sepsis 3 criteria, and for the vasopressor cohort over the 12 hours before and after norepinephrine initiation. Fluids included crystalloids (0.9% sodium chloride, PlasmaLyte, Lactated Ringer’s) and albumin. In the vasopressor cohort, use of second line agents included epinephrine, vasopressin, and phenylephrine, and adjunctive therapies such as corticosteroids were collected. Patients receiving <30 mL/kg fluid resuscitation were screened but excluded from the final analysis (Supplemental Figure 1).

### Outcomes

The primary outcome was mortality for 7 days following ICU admission. Secondary outcomes included mortality in 1-day, persistent dysfunction of end-organ systems (i.e., renal, neurological, respiratory) at day 28 and comparison of survival between groups. The criteria for organ failure are described in Supplementary Section 2 and Supplementary Table 1. Subgroup analysis within the fluids-plus-vasopressor cohort assessed whether exposure to second-line vasopressors or adjunctive therapies was associated with differences in MSE profiles or clinical outcomes.

### Data Abstraction

Physiological waveform data were collected using General Electric (GE) bedside monitors and the BedMaster system (Excel Medical Electronics, Jupiter, FL, USA). High-resolution ECG waveforms were extracted from the bedside monitors and sampled at 240 Hz. MSE features were derived using the PhysioNet Cardiovascular Signal Toolbox.^15^ Preprocessing included ECG signal filtering and R-peak detection, followed by extraction of heart rate variability (HRV) features spanning time-domain, frequency-domain, nonlinear metrics, and multiscale sample entropy across scales 1–20 to characterize short-to long-term autonomic variability (Supplemental Figure 2). These features were subsequently used as inputs to machine learning models for autonomic phenotyping and outcome prediction. The features were calculated using 300 second windows with a 30-millisecond overlap. For each patient, the baseline segment was defined as the 1-hour period beginning 24 hours after ICU admission; if unavailable, the nearest 1-hour segment within the preceding 24 hours was used. HRV and MSE values within this segment were aggregated using the mean to generate a single baseline MSE profile for each patient.

## Statistical Analysis

To minimize confounding, propensity score matching using a 1:1 nearest neighbor approach was performed based on age, sex and clinical illness severity scores (APACHE II, SOFA) at 24 hours post ICU-admission. Kaplan–Meier analysis compared survival between the fluids-only and fluids-plus-vasopressor cohorts, with differences assessed using the log-rank test and odds ratios. Outcomes within each cohort were further stratified by survivor status. Statistical comparisons were performed using Student’s t-test for normally distributed variables and the Mann–Whitney U test for non-parametric data. To account for multiple comparisons, p-values were adjusted using the Bonferroni correction. Data visualization was performed using Python and R, including line plots, Principal Component Analysis (PCA), Uniform Manifold Approximation and Projection (UMAP) embeddings, and violin plots.

### Machine Learning and Clustering Analysis

Supervised machine learning algorithms including logistic regression, eXtreme Gradient Boosting (XGBoost), random forest, and gradient boosting were evaluated to predict the primary and secondary outcomes. For the predictive modeling, the MSE complexity scales were further subdivided into seven aggregated features representing progressively increasing temporal resolution: Ultra Short Term (scales 1–2), Very Short Term (scales 3–4), Short Term (scales 5–6), Intermediate Term (scales 7–9), Long Term (scales 10–12), Very Long Term (scales 13–15), and Ultra Long Term (scales 16–20). Each feature was defined as the mean MSE value across its constituent scales. Three predictive models were developed for comparative evaluation: (1) A model including only entropy-derived features (MSE), (2) a model incorporating entropy features along with demographic and vital sign variables extracted from the EMR (MSE-DV), (3) HRV model including features from time-domain, frequency-domain and nonlinear-domain. Time-domain measures summarized central tendency, dispersion, and distributional properties of NN intervals (NNmean, NNmedian, NNmode, NNvariance, NNiQR, NNskew, NNkurt), while standard variability indices (SDNN, RMSSD, pNN50) reflected overall and parasympathetic-mediated variability. Frequency-domain features quantified spectral power in ultra-low, very-low, low, and high-frequency bands (ULF, VLF, LF, HF), total power, and the LF/HF ratio, capturing autonomic modulation across time scales. Nonlinear features derived from Poincaré plots (SD1, SD2, SD1/SD2) and acceleration and deceleration capacities (AC, DC) characterized short- and long-term heart rate dynamics (4) a model constructed with established severity of illness indices (SOAP). Demographic and vital sign variables included race, sex, age, temperature, mean arterial pressure (MAP), diastolic blood pressure (DBP), systolic blood pressure (SBP), and unassisted respiratory rate. Severity of illness indices included SOFA and APACHE II scores calculated over the same 24-hour timeframe as the entropy and vital sign data. Hyperparameter tuning was performed using a grid search strategy.^16^ To preserve class proportions and quantify model uncertainty, stratified bootstrapping was performed with 1000 iterations. Predictors were standardized prior to model training, and classification thresholds were chosen to achieve 80% specificity, ensuring a clinically meaningful balance of sensitivity and specificity. Area Under the Receiver Operating Characteristic Curve (AUROC) was the primary metric evaluated for predictive performance. Additional performance indices included sensitivity, specificity, positive predictive value (PPV), and negative predictive value (NPV). Feature importance for mortality prediction and organ dysfunction was quantified using SHapley Additive exPlanations (SHAP) values to facilitate clinical understanding of model predictions.^17^ The dataset was randomly split into a training set (80%) and a testing set (20%), and model generalizability was evaluated using five-fold cross-validation. Calibration was evaluated using reliability diagrams and the Brier score. SHAP-based interpretability was applied. The methodology for organ dysfunction prediction and physiological phenotyping is described in detail in Supplementary Section 2.

## Results

A total of 2,374 patients with sepsis or septic shock were screened for inclusion. Of these, 298 patients were excluded for receiving less than 30 mL/kg of intravenous fluids, resulting in 2,079 eligible patients. Of these, 479 patients who did not receive norepinephrine were classified as the fluids-only cohort. The remaining 1,600 patients received ≥30 mL/kg of intravenous fluids plus norepinephrine and formed the fluids-plus-vasopressor cohort. Propensity score matching produced a final matched cohort of 958 patients for comparative analysis (479 matched pairs). Figure 1 provides a CONSORT diagram for this process and Supplemental Table 2 shows the effect sizes for matching.

**Figure 1.**
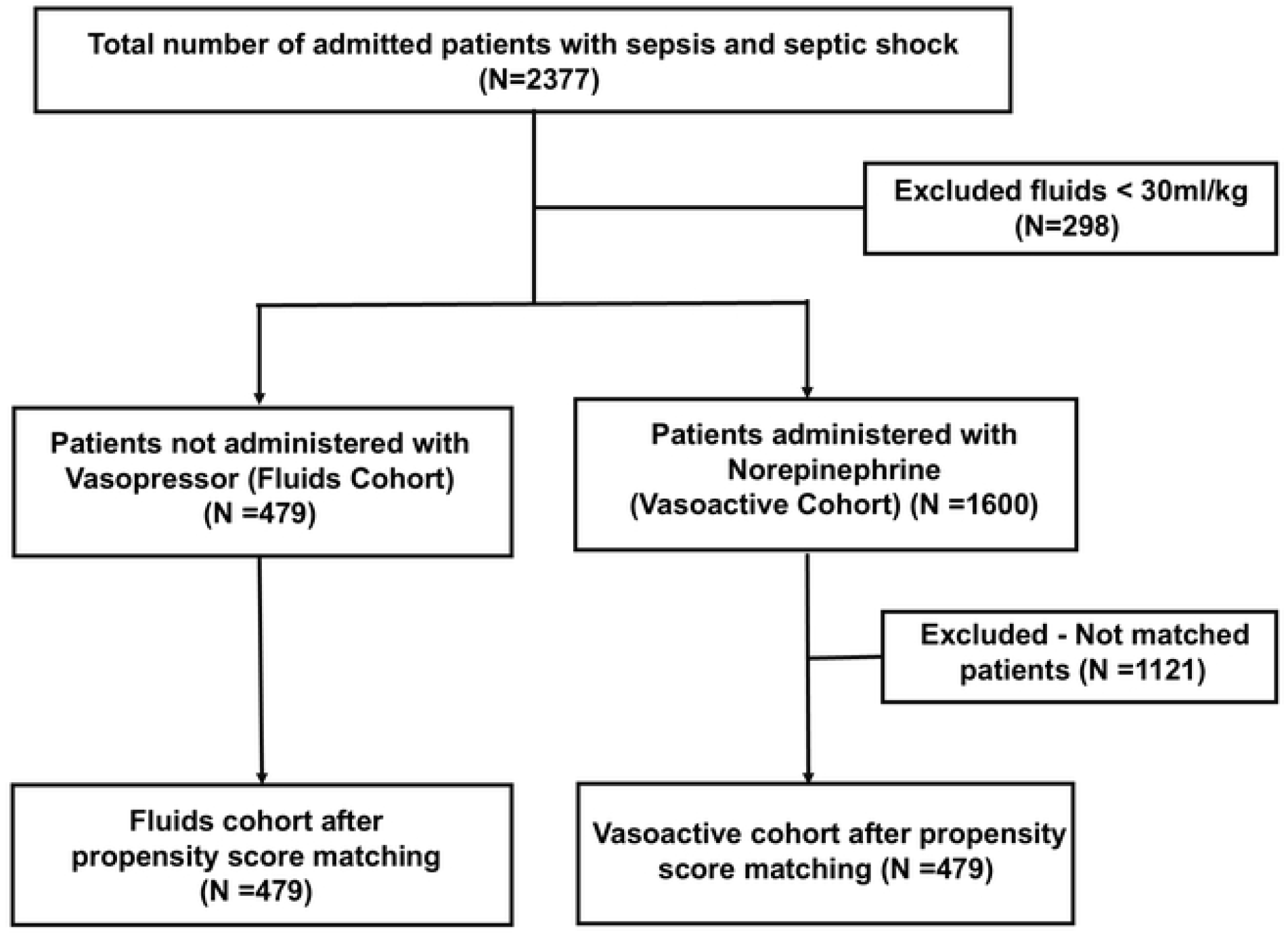
Consort diagram illustrating the patient selection process. The mean age of the matched cohort was 59.7 years (SD 17.5). A higher proportion of younger patients (18–40 years) were present in the fluids-only cohort compared with the fluids-plus-vasopressor group (19% vs 12.1%, p < 0.001). Sex and racial distributions were similar between cohorts, with most patients identified as Caucasian/White (50.6%) or African American/Black (41.3%). Illness severity, as measured by SOFA or APACHE II scores at 24 hours, did not differ between cohorts (Table 1). Additional baseline patient characteristics are reported in Supplemental Table 3.

**Table 1:** Demographics characteristics for fluids-only and fluid-plus-vasopressor cohorts.

|  |  | Overall | Fluids-only | Fluids-plus-vasopressor | P-Value |
| --- | --- | --- | --- | --- | --- |
|  | n | 958 | 479 | 479 |  |
| Age, mean (SD) |  | 59.7 (17.5) | 59.3 (19.3) | 60.2 (15.5) | 0.397 |
| Age Group, n (%) | 18-40 yrs. | 149 (15.6) | 91 (19.0) | 58 (12.1) | <0.001 |
|  | 41-60 yrs. | 288 (30.1) | 134 (28.0) | 154 (32.2) |  |
|  | 61-80 yrs. | 401 (41.9) | 179 (37.4) | 222 (46.3) |  |
|  | 81+ yrs. | 120 (12.5) | 75 (15.7) | 45 (9.4) |  |
| Sex, n (%) | Female | 475 (49.6) | 248 (51.8) | 227 (47.4) | 0.196 |
|  | Male | 483 (50.4) | 231 (48.2) | 252 (52.6) |  |
| Race, n (%) | Caucasian or White | 485 (50.6) | 231 (48.2) | 254 (53.0) | 0.134 |
|  | African American or Black | 396 (41.3) | 216 (45.1) | 180 (37.6) |  |
|  | Asian | 34 (3.5) | 16 (3.3) | 18 (3.8) |  |
|  | Native Hawaiian or Other Pacific Islander | 3 (0.3) | 2 (0.4) | 1 (0.2) |  |
|  | Multiple | 6 (0.6) | 2 (0.4) | 4 (0.8) |  |
|  | Unknown, Unavailable or Unreported | 34 (3.5) | 12 (2.5) | 22 (4.6) |  |
| Unit, n (%) | Medical ICU | 409 (42.7) | 263 (54.9) | 146 (30.5) | <0.001 |
|  | Surgical ICU | 252 (26.3) | 113 (23.6) | 139 (29.0) |  |
|  | Cardiothoracic ICU | 125 (13.0) | 26 (5.4) | 99 (20.7) |  |
|  | Neuro ICU | 121 (12.6) | 56 (11.7) | 65 (13.6) |  |
|  | Cardiac Care Unit | 51 (5.3) | 21 (4.4) | 30 (6.3) |  |
| SOFA (max), mean (SD) |  | 4.3 (2.8) | 4.5 (2.2) | 4.2 (3.3) | 0.148 |
| APACHEII Score, mean (SD) |  | 12.2 (6.3) | 12.5 (5.2) | 11.9 (7.1) | 0.121 |

Figure 2 illustrates the relationship between multiscale entropy (MSE) features and 1-day mortality. In the fluids-plus-vasopressor cohort, patients who expired within 24 hours exhibited significantly lower entropy across all temporal scales compared to survivors (Figure 2A–B), with large effect sizes observed in mid-range and long-term complexity domains. Dimensionality reduction using PCA and unsupervised k-means clustering revealed two distinct physiological clusters, with one cluster enriched for early mortality (Figure 2C). Although survivors and non-survivors overlap, regions of higher mortality density emerge, and unsupervised clusters capture latent states that partially align with clinical outcome, supporting the prognostic value of entropy-based metrics. When comparing fluids-plus-vasopressor expired patients to fluids-only survivors, similar reductions in entropy were observed across short-to long-term scales (Figure 2D–E), again demonstrating large effect sizes. Clustering analysis of this combined cohort revealed that patients with reduced MSE clustered distinctly, regardless of treatment assignment (Figure 2F). Logistic regression analysis identified lower mid-range and long-term entropy as significant risk factors for early mortality, whereas preserved complexity in these domains was associated with reduced risk (Figure 2G). Kaplan–Meier curves, stratified by treatment group, further demonstrated a pronounced divergence in survival probability within the first 24 hours, with the fluids-plus-vasopressor cohort showing significantly higher early mortality (Figure 2H).

**Figure 2.**
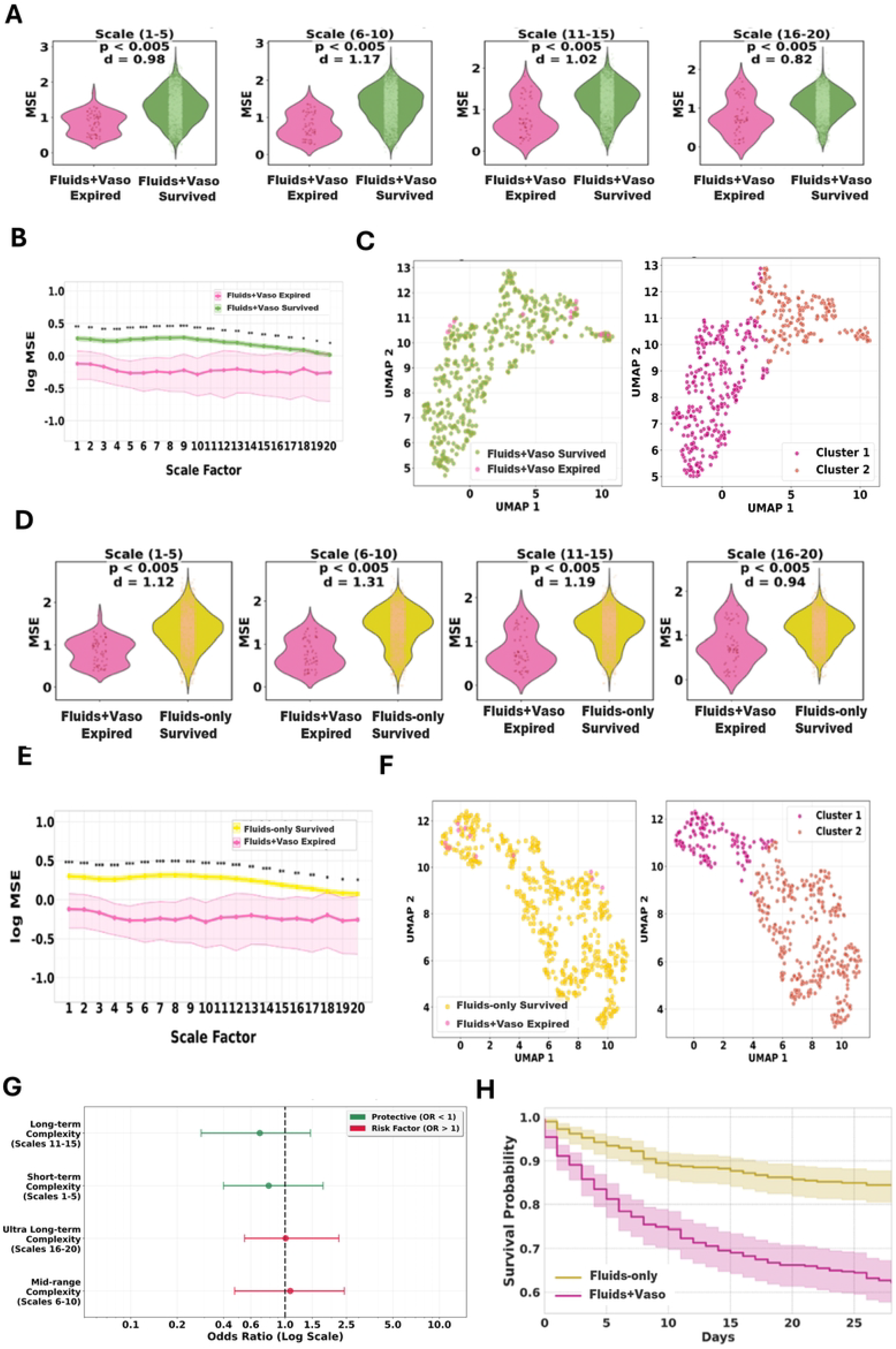
MSE-based stratification of short-term (1-day) mortality outcomes and phenotypes in sepsis. (A) Violin plots showing significantly lower MSE values across all grouped scale ranges (short-term: 1–5, mid-range: 6–10, long-term: 11–15, ultra long-term: 16–20) in expired (n=12) vs. survived (n=467) patients within the fluids-plus-vasopressor cohort. Cohen’s d indicates medium to large effect sizes (B) Mean log-transformed MSE values across all 20 scales, showing consistent reductions in complexity among vasoactive expired patients compared to survivors. (C) UMAP projection of MSE features showing spatial separation of fluids-plus-vasopressor survivors and expired patients (left), and unsupervised k-means clustering identifying two physiologic clusters (right). (D) Violin plots comparing fluids-plus-vasopressor expired patients (n=12) and fluids-only survivors (n= 468), revealing significantly lower MSE across all scale ranges, with large effect sizes (E) Line plot of log MSE values across scales demonstrating reduced complexity in fluids-plus-vasopressor expired patients relative to fluids-only survivors. (F) UMAP projection of MSE features in the fluids vs. vasoactive group comparison, with left panel showing outcome overlay and right panel showing unsupervised k-means cluster assignment. (G) Forest plot showing odds ratios (log scale) from multivariable logistic regression indicating that reductions in mid-range and long-term entropy are associated with increased risk of mortality, while preserved complexity in long-term scales appears protective. (H) Kaplan–Meier survival curves showing significantly lower survival probability over 28 days in the fluids-plus-vasopressor cohort compared to the fluids-only cohort.

The performance of the short-term (7-day) mortality prediction models for the fluids-plus-vasopressor cohort and the pooled cohort (fluids-only and fluids-plus-vasopressor patients combined) is presented in Table 2. MSE only demonstrated good discrimination without any demographic or vitals features. In the vasoactive cohort it achieved an AUROC of 0.77 (0.63–0.88) substantially higher than SOAP (0.64 [0.45–0.82]) with specificity 0.80 (0.75–0.86) at the prespecified operating point and a high NPV of 0.91 (0.85–0.90). Adding demographics/vitals (MSE-DV) yielded a AUROC gain of 7% (0.84 [0.73–0.92]), the MSE-only model retained clinically useful performance. In the combined vasoactive + fluids cohort analysis, discrimination was lower overall, but the pattern persisted: MSE-DV 0.74 (0.61–0.86), MSE 0.72 (0.58–0.83) and SOAP 0.60 (0.45–0.75). Sensitivity/specificity pairs were 0.46 / 0.75 for MSE-DV, 0.30 / 0.86 for MSE, and 0.30 / 0.80 for SOAP. PPV was modest (0.10–0.26) while NPV remained high (0.91–0.95), consistent with utility for ruling out short-term mortality in this setting. Across algorithms (Random Forest, XGBoost, Gradient Boosting, Logistic Regression), Random Forest consistently provided the highest discrimination and most favorable sensitivity specificity trade-offs; therefore, Random Forest was used as the primary classifier for all model comparisons reported. These findings indicate that multiscale entropy features alone capture a strong mortality signal, with added variables providing incremental improvement. Supplemental Table 4-7 shows the performance of machine learning algorithms including XGBoost, Gradient Boosting, SVM and logistic regression. Calibration results have been described in Supplemental Section 3. Across both fluids-plus-vasopressor and pooled cohorts, MSE models consistently outperformed HRV models. In the fluids-plus-vasopressor cohort, MSE demonstrated higher discrimination than HRV (AUROC 0.77 vs 0.75) with improved sensitivity and comparable specificity. Similar trends were observed in the pooled cohort, where MSE model maintained superior AUROC compared with HRV (0.72–0.74 vs 0.66). Notably, the MSE model used only seven grouped entropy scales, yielding a parsimonious representation that preserved high discrimination and negative predictive value, in contrast to HRV models with broader time-domain, frequency-domain and nonlinear-domain feature sets.

**Table 2:** Performance of short-term (7-day) mortality prediction model for vasoactive cohort.

| Model | AUROC | Sensitivity | Specificity | PPV | NPV |
| --- | --- | --- | --- | --- | --- |
| MSE<br>(Fluids-plus<br>vasopressor) | 0.77<br>(0.63, 0.88) | 0.45<br>(0.36, 0.63) | 0.80<br>(0.75, 0.86) | 0.22<br>(0.12, 0.32) | 0.91<br>(0.85, 0.90) |
| MSE-DV<br>(Fluids-plus-vasopressor) | 0.84<br>(0.73, 0.92) | 0.54<br>(0.42, 0.68) | 0.80<br>(0.75, 0.86) | 0.26<br>(0.16, 0.37) | 0.93<br>(0.86, 0.92) |
| HRV<br>(Fluids-plus-vasopressor) | 0.75<br>(0.59, 0.87) | 0.36<br>(0.21, 0.52) | 0.83<br>(0.78, 0.88) | 0.222<br>[0.10, 0.32]) | 0.910<br>[0.84, 0.90]) |
| SOAP<br>(Fluids-plus-vasopressor) | 0.64<br>(0.45, 0.82) | 0.45<br>(0.36, 0.63) | 0.78<br>(0.74, 0.86) | 0.21<br>(0.11, 0.31) | 0.91<br>(0.85-0.90) |
| MSE<br>(Pooled) | 0.72<br>(0.58, 0.83) | 0.30<br>(0.19, 0.52) | 0.86<br>(0.83, 0.90) | 0.13<br>(0.05, 0.20) | 0.94<br>(0.91, 0.94) |
| MSE-DV<br>(Pooled) | 0.74<br>(0.61, 0.86) | 0.46<br>(0.38, 0.66) | 0.75<br>(0.72, 0.82) | 0.12<br>(0.04, 0.16) | 0.95<br>(0.91, 0.95) |
| HRV<br>(Pooled) | 0.66<br>(0.53, 0.77) | 0.33<br>(0.19, 0.52) | 0.8101<br>(0.78, 0.87) | 0.10<br>(0.04, 0.14) | 0.94<br>(0.90, 0.94) |
| SOAP<br>(Pooled) | 0.60<br>(0.45, 0.75) | 0.30<br>(0.19, 0.52) | 0.80<br>(0.77, 0.86) | 0.10<br>(0.03, 0.14) | 0.94<br>(0.90, 0.93) |

Table 3 summarizes 28-day organ dysfunction performance in the fluids-plus-vasopressor cohort using a Random Forest with the MSE-DV feature set. Discrimination was highest for neurological dysfunction (AUROC - 0.79), followed by renal failure (0.75) and respiratory failure (0.71). NPV were consistently high (0.84 – 0.94) and PPV low (0.15 – 0.45), indicating greater utility for ruling out 28-day organ dysfunction. Performance for the combined vasoactive + fluids cohort using the Random Forest MSE-DV model is shown in Supplemental Table 8. Discrimination was attenuated, with AUROCs only 0.55–0.63 across organ outcomes. This reduction in performance reflects lower prevalence of organ failure outcomes and greater clinical heterogeneity when the cohorts are combined.

**Table 3:** Performance of 28-day organ dysfunction prediction model (MSE-DV) for fluids-plus-vasopressor cohort.

| Organ Dysfunction | AUROC | Sensitivity | Specificity | PPV | NPV |
| --- | --- | --- | --- | --- | --- |
| Renal Failure | 0.75<br>(0.60, 0.86) | 0.42<br>(0.31, 0.59) | 0.78<br>(0.73, 0.84) | 0.25<br>(0.13, 0.34) | 0.88<br>(0.82, 0.88) |
| Neurological Dysfunction | 0.79<br>(0.69, 0.88) | 0.50<br>(0.36, 0.66) | 0.82<br>(0.76, 0.87) | 0.45<br>(0.38, 0.59) | 0.84<br>(0.78, 0.87) |
| Respiratory Failure | 0.71<br>(0.54, 0.86) | 0.42<br>(0.33, 0.60) | 0.82<br>(0.77, 0.87) | 0.15<br>(0.07, 0.22) | 0.94<br>(0.88, 0.92) |

In the fluids-plus-vasopressor cohort (Figure 3A–C), MSE only features (A) showed that short-scale complexity was the most important feature followed by longer scales. After adding demographics/vitals (Figure 3B), the importance profile shifted toward temperature and MAP, with age and respiratory rate contributing, while long-scale MSE features remained among the top predictors. Consistent with these feature attributions, the ROC curves (Figure 3C) show MSE-DV performing best, MSE-only retaining strong discrimination, and SOAP lagging. In the pooled (fluids-only and fluids-plus-vasopressor combined) cohort (Figure 3D–F), the MSE-only model (Figure 3D) emphasized long-range complexity alongside short-scale features, whereas adding demographics/vitals (Figure 3E) shifted importance toward gender, temperature, and race, with long-scale MSE still contributory. ROC curves for the combined cohort (Figure 3F) show modest but consistent performance MSE-DV slightly higher than MSE-only, both outperforming SOAP for 7-day mortality prediction.

**Figure 3.**
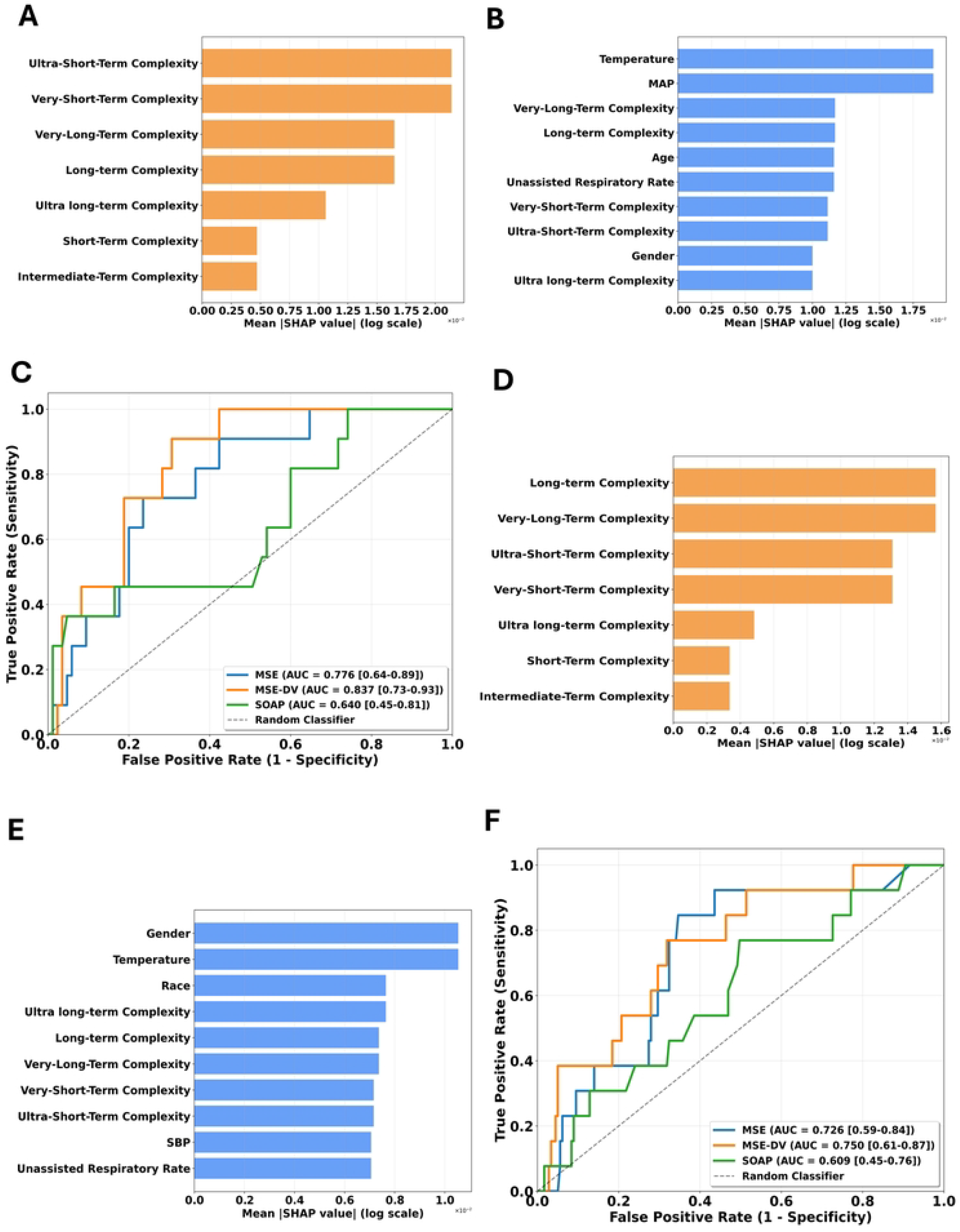
Feature Importance and Random Forest Model Performance for short term (7-day) mortality prediction. Fluids-plus-vasopressor cohort (A–C): (A) SHapley Additive exPlanations (SHAP) mean absolute values for the MSE-only (B) With MSE + demographic/vital-sign features (MSE-DV) (C) ROC curves for MSE-DV model showed superior performance compared to clinical scoring model – SOFA+APACHE ii (SOAP). MSE-only model showed acceptable performance. Pooled cohort (D–F) (fluids-only and fluids-plus-vasopressor combined: (D) The important SHAP features for MSE only model for combined cohort are long and very-long-term complexities. (E) In MSE-DV, demographic/vital-sign features gain prominence with contributions from long-scale MSE. (F) ROC curves show superior discrimination for MSE-DV model compared to clinical scoring model (SOAP).

Figure 4. shows MSE changes across vasopressor escalation and corticosteroid exposure. Supplemental Table 9 shows the distribution of vasopressor and steroid therapies. Across fluids-plus-vasopressor treated patients, escalation beyond NE was associated with lower MSE, most notably at mid and long-range scales. In second-line therapy, the distribution of MSE shifted downward for EPI compared with NE alone, indicating a global loss of complexity rather than outlier effects; across agents the decrement ranked EPI > VP >PE. Third-line combinations produced the deepest, near-uniform reductions across scales. Corticosteroid exposure was linked to additional complexity loss, most evident with NE + PE + HC. Increasing therapeutic intensity and steroid use aligned with a progressive, system-wide decline in MSE, indicative of worsening shock and impaired physiological resilience.

**Figure 4.**
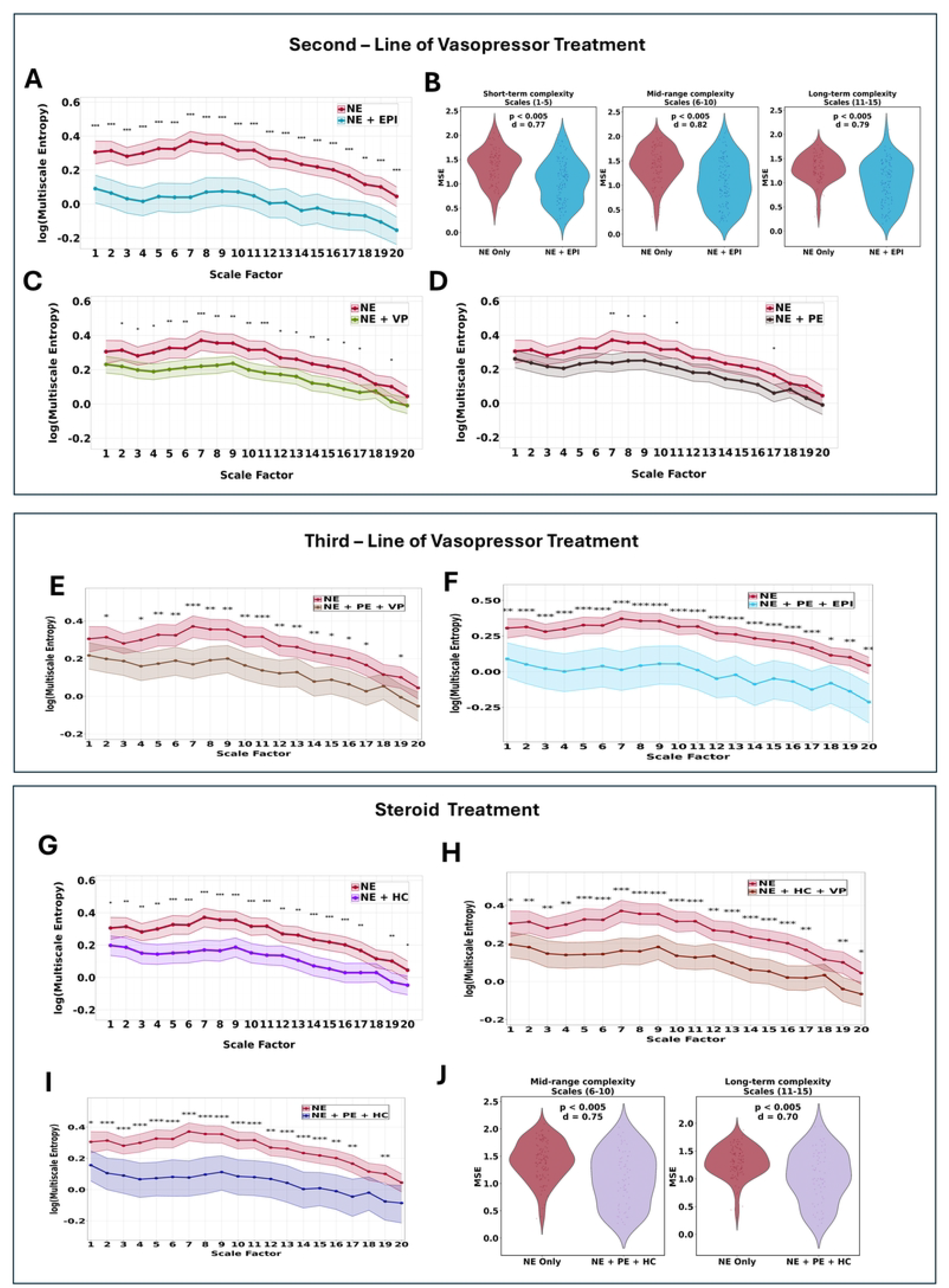
Multiscale Entropy (MSE) reveals physiological complexity differences across fluids-plus-vasopressor treatment subgroups. (A) Comparison between norepinephrine (NE) only and NE + epinephrine (EPI) subgroups shows significantly reduced entropy across all scale ranges in the NE + EPI group. (B) Violin plots demonstrate moderate to large effect sizes, indicating greater autonomic dysregulation in patients requiring epinephrine. (C-D) Comparison plots of NE versus combination of NE and adjunct vasopressors vasopressin (VP), phenylephrine (PE) shows reduced lower complexity. The most pronounced scales are (7–11) for VP and scale 7 for PE. (E-F) Third line of vasopressor treatment show consistently lower log(MSE) than NE alone across scales (1–20), most pronounced at mid–long scales (6–15). (G) Log (MSE) across scales 1–20 for NE compared to NE+ hydrocortisone (HC) shows consistently lower complexity, most evident at scales (5-16). (H) Log (MSE) for NE vs NE+HC+VP. Corticosteroid + vasopressin further reduces complexity compared with NE alone. (I) Log (MSE) for NE vs NE+PE+HC. Corticosteroid + phenylephrine group shows a broad downward shift in complexity across scales, with the largest gaps at mid–long ranges. (J) Violin plots of mid-range (6–10) and long-range (11–15) complexity: NE+PE+HC distributions are shifted downward compared to NE (p<0.005; large effects), confirming reduced complexity with steroid multiagent escalation. Statistical significance (p < 0.05 to p < 0.0001) across multiple scales supports the discriminative utility of MSE for stratifying patients by vasopressor escalation patterns.

## Discussion

In this study, we demonstrate that MSE derived from high resolution physiological waveform data within the first 24 hours of ICU admission provides meaningful prognostic value in patients with sepsis and septic shock. Our findings indicate that baseline entropy features are predictive of short-term (7-day) mortality, with AUROC values up to 0.84 in the fluids-plus-vasopressor cohort and consistently high NPVs. These findings suggest potential utility of entropy-based metrics for ruling out early mortality, a clinically important use case given the need to identify lower-risk patients who may not require immediate escalation of monitoring or intervention. The cohort’s racial diversity and inclusion of patients from multiple ICU types support the generalizability of our findings across heterogenous patient populations and clinical environments.

Across all evaluated models, entropy features alone, spanning multiple temporal domains, ranging from ultra-short to ultra-long-term complexity, captured a strong mortality signal. The addition of demographic and vital sign variables yielded only incremental improvements. This highlights the ability of MSE to capture physiological regulation across multiple time horizons, reflect latent autonomic dysfunction, and independently convey prognostic information. When compared against traditional severity of illness indices such as SOFA and APACHE II, MSE consistently demonstrated superior discrimination. Whereas these severity of illness scores summarize organ dysfunction at discrete time points, entropy measures directly quantify the intrinsic variability and complexity of physiologic systems, offering a more dynamic representation of underlying autonomic and cardiovascular regulation.^18^ This mechanistic distinction likely accounts for the enhanced predictive ability of MSE in identifying patients at risk of early deterioration.

SHAP interpretability analysis further confirmed the centrality of entropy features. In the fluids-plus-vasopressor cohort, very-short-term and ultra-short-term complexity were the most influential predictors, followed by long-term and very-long-term complexity. This pattern suggests that both rapid fluctuations and slower scale complexity loss contribute to early mortality risk in patients requiring vasopressor support. In contrast, in the pooled cohort demographic and physiological variables (sex, temperature, race) gained relative prominence while long-term and very-long-term complexity remained important. This shift is consistent with the lower AUROC values in the pooled cohort, implying that entropy-derived signatures are most discriminative in high-risk patients with greater physiological stress, while background demographic and vital sign variation becomes more explanatory in broader populations. Interestingly, age, traditionally an important risk marker, demonstrated relatively low feature importance in both the fluids-plus-vasopressor and pooled cohorts, particularly when compared to MSE-derived complexity measures and vital signs. This may reflect the ability of MSE to capture real-time physiological resilience and autonomic regulation, which are more directly linked to short-term outcomes than chronological age.

Long-term (scales 11–18) entropy domains emerged as especially informative in stratifying patients by both treatment intensity and clinical trajectory. These scales demonstrated substantial reductions in patients requiring second and third line therapies beyond norepinephrine, including epinephrine, vasopressin, phenylephrine, and corticosteroids. Such therapies are typically reserved for more refractory shock states,^19^ and the associated declines in entropy suggest deeper autonomic impairment and reduced homeostatic capacity.^20^ In particular, the observed reduction in MSE across all temporal scales in patients receiving epinephrine suggests profound autonomic dysregulation and loss of physiological complexity. Epinephrine’s potent adrenergic stimulation may suppress intrinsic HRV, while its use often reflects a more severe shock state, potentially involving sepsis-induced myocardial dysfunction, a condition known to impair cardiac autonomic regulation and baroreflex sensitivity, contributing to entropy loss. Overall, MSE at these longer time scales likely captures cumulative burden on cardiovascular reflexes and neuroendocrine signaling, providing insight into latent autonomic exhaustion and loss of adaptability during septic progression.^21,22^ Specifically, this pattern was seen in patients who experienced acute kidney injury and respiratory failure and neurological decline indicating early loss of physiological complexity. This supports the hypothesis that reduced variability across multiple temporal scales reflects autonomic dysfunction, signaling a shift toward rigid, less adaptive physiology and impaired system resilience mechanisms that are central to sepsis pathophysiology.^23–25^

Conventional HRV measures including time, frequency, and non-linear domain indices provide valuable insights into autonomic function but primarily reflect static, single-point assessments.^26^ These metrics often fail to capture the evolving physiological adaptations and system-wide interactions that occur in critical illness. In contrast, multiscale entropy (MSE) provides a dynamic, scale-sensitive assessment of signal complexity by evaluating entropy across multiple temporal resolutions.^27^ This makes MSE particularly well-suited to capture the temporal disorganization that occurs during systemic stress responses, including those driven by dysregulation of the neuroendocrine-immune axis.^28^ The hypothalamic-pituitary-adrenal (HPA) axis, autonomic nervous system, baroreflex sensitivity, circadian rhythms, and inflammatory networks critical for maintaining homeostasis during a robust stressor like septic shock operate over distinct time scales, and MSE reflects this multi-time-scale coordination, offering a more integrated measure of physiological resilience.^29^ As sepsis progresses, the breakdown of this coordination manifests as reduced entropy across short- and long-range scales an effect that MSE can detect more sensitively than traditional HRV indices.^30,31^ In summary, our findings support MSE as a non-invasive, interpretable biomarker that captures early physiological deterioration in sepsis. Integrating entropy-based analytics into ICU decision-support systems may help identify high-risk patients who require closer monitoring or earlier intervention.

This study has limitations. As a single center study, findings may be influenced by differences in ICU types and care practices, potentially affecting both treatment decisions and physiological monitoring and thus confounding entropy based comparisons. Our primary cross-sectional analysis compared patients receiving fluids-plus-vasopressors with those receiving fluids alone; although propensity score matching was applied to adjust for baseline illness severity, residual confounding from unmeasured factors such as autonomic tone and comorbidity burden are possible. Vasopressor timing and dosing were not controlled for, which may further complicate interpretation of entropy changes. Mechanical ventilation and sedation, known to suppress autonomic variability, were not explicitly modeled and may influence MSE values. Finally, patients with pacemakers or persistent atrial fibrillation, conditions that markedly alter HRV and entropy, may not have been fully excluded, potentially affecting observed variability patterns. Future work should incorporate ventilator parameters and sedation depth to better isolate intrinsic autonomic signals.

## Conclusion

MSE derived from ICU ECG data was associated with short-term mortality and longer-term organ dysfunction in patients with sepsis and septic shock. Entropy-based models demonstrated higher discrimination than models based on conventional severity of illness scores. These findings support the potential role of MSE in early risk stratification and motivate further evaluation in larger, multicenter cohorts.

## Author Contributions

Conceptualization, Investigation, Methodology: P.K., R.K., A.S., B.M.

Data Curation: P.K., A.G.

Formal Analysis: P.K.

Validation, Visualization: P.K., R.K., A.S., B.M.

Study Design: P.K., R.K., A.S., B.M.

Supervision and Funding Acquisition: R.K.

Writing – Original Draft: P.K.

Writing – Review and Editing: P.K., R.K., A.S., B.M., A.A., M.P., P.U., A.G., T.C., A.L.H., A.E.

## Data availability statement

The data cannot be made publicly available upon publication due to legal restrictions preventing unrestricted public distribution. The data that supports the findings of this study are available upon reasonable request from the authors.

---

**Supplemental Figure 1.**
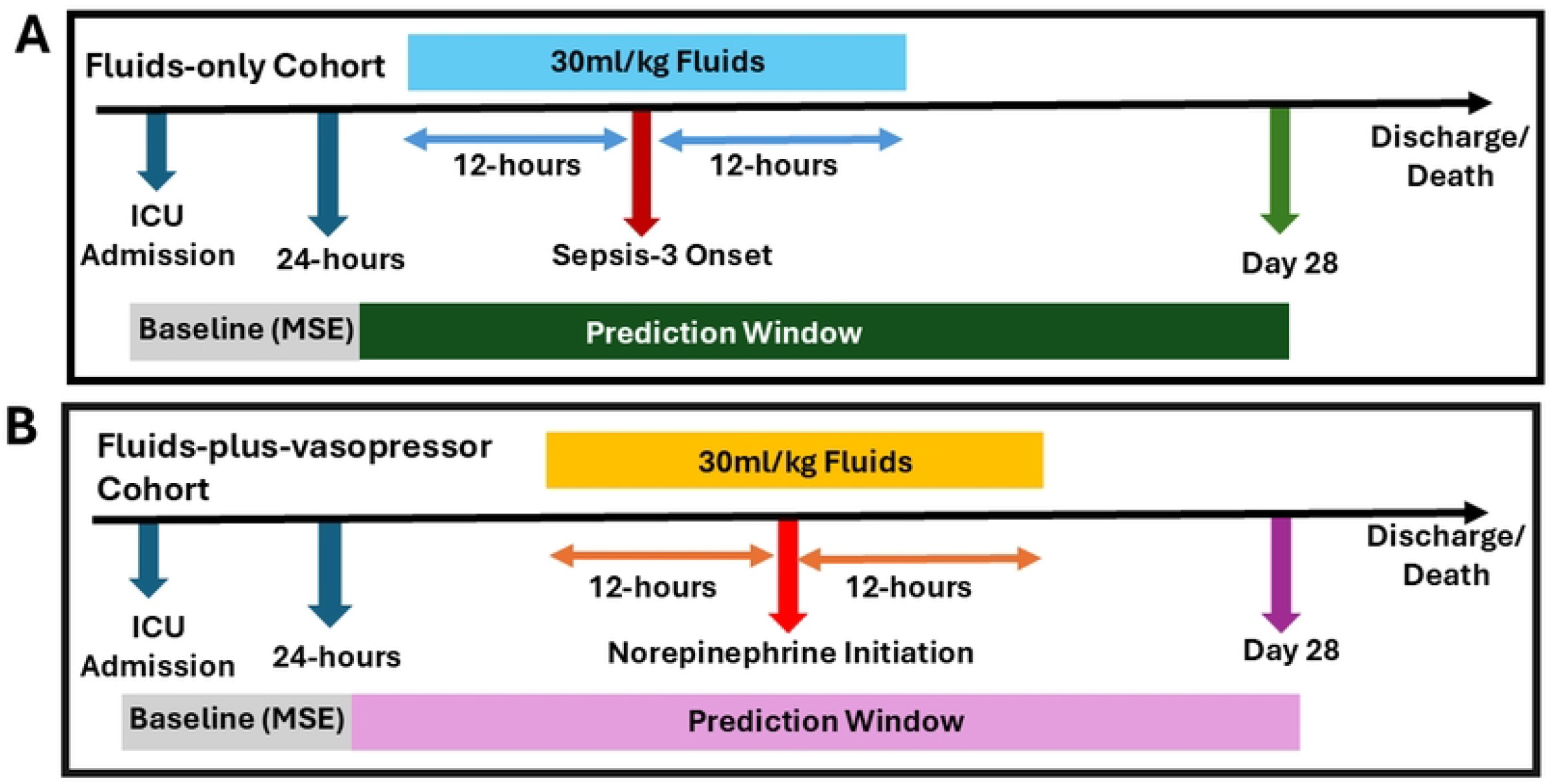

**Supplemental Figure 2.**
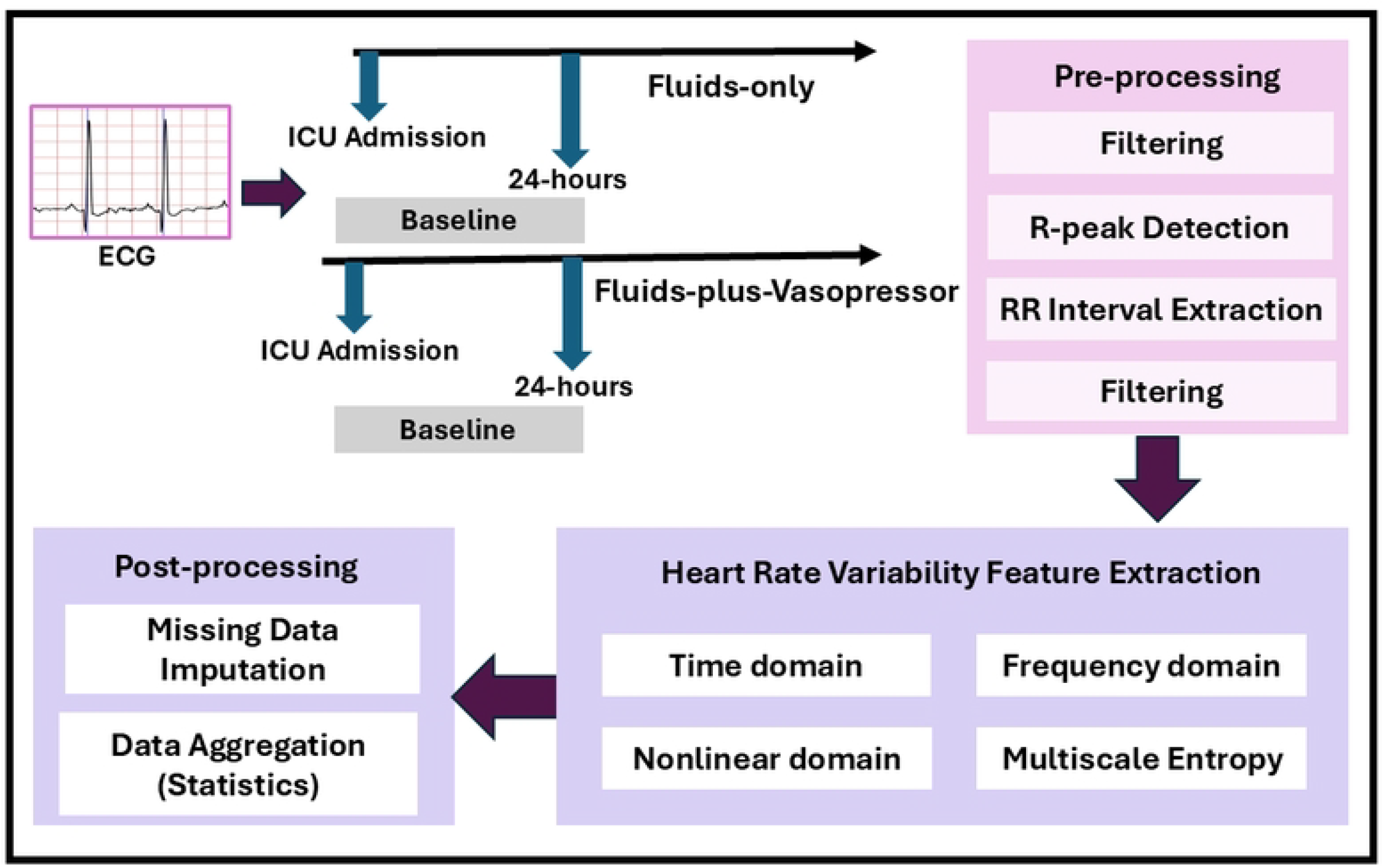

**Supplemental Figure 3.**
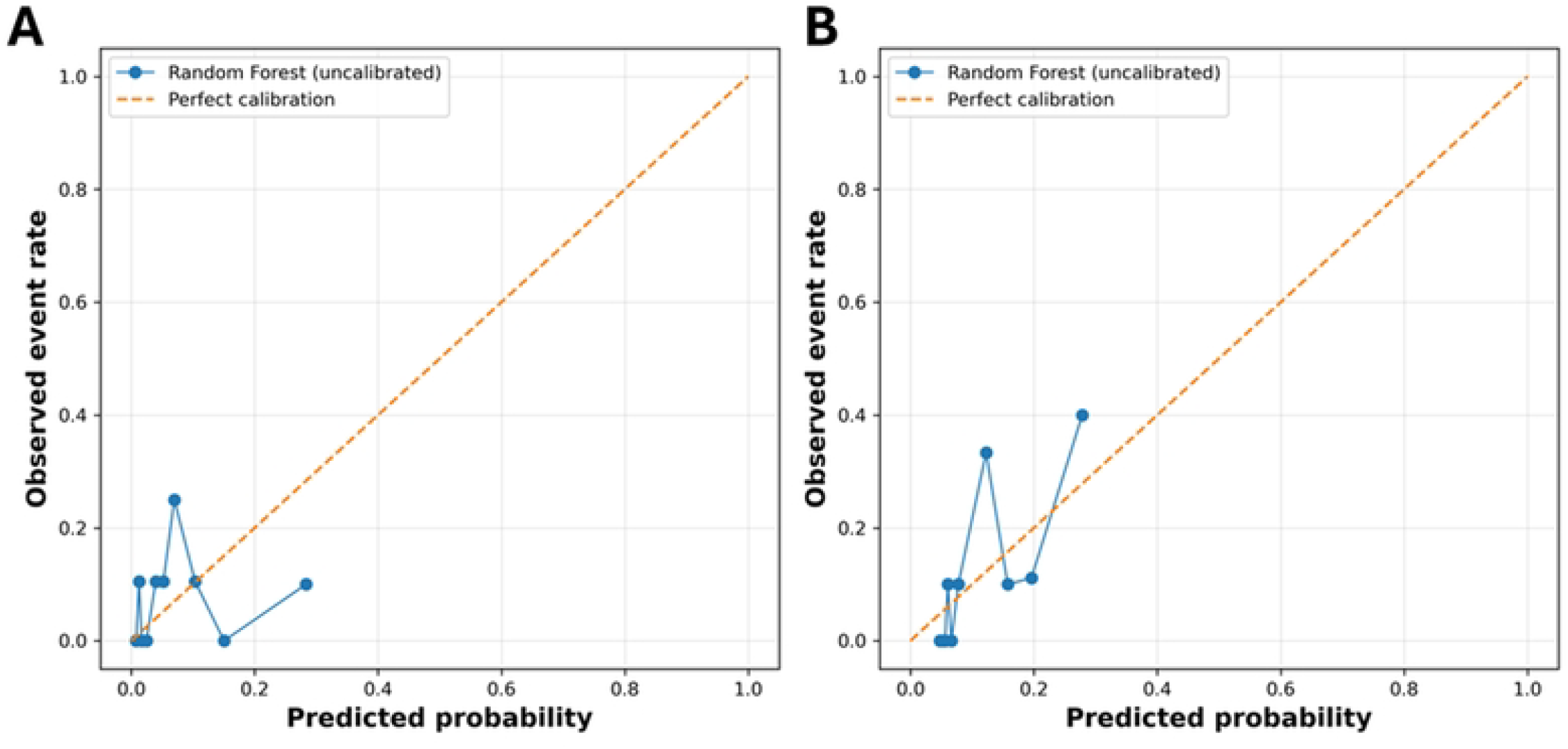

